# Socio-economic inequalities and cancer mortality among adolescents and young adults between 2000 and 2023

**DOI:** 10.64898/2026.11.02.26367080

**Authors:** Essi Ruth Kossi, Jacques Demongeot, Leila Gofti Laroche

## Abstract

Cancer among adolescents and young adults (AYAs) has been a growing public health concern in France since 2009, a priority reaffirmed in the new 2026-2030 roadmap of the Ten-Year Cancer Control Strategy. This issue is also generating increasing interest internationally, as evidenced by the emergence of scientific conferences dedicated to this population. However, the socioeconomic inequalities associated with cancer mortality among AYAs remain insufficiently documented internationally. The objective of this study is to analyze the socioeconomic determinants associated with cancer mortality among AYAs. Epidemiological data on cancer patients aged 15 to 39 are available for 37 countries for 2000 to 2023, these data being drawn from the Global Burden of Disease (GBD 2023) databases. For this study, they were supplemented by socioeconomic indicators from the United Nations Development Program and the World Bank. Descriptive analyses, correlations, principal component analysis, classification methods, time-series trend analyses (EAPC and Joinpoint), and panel data models have been performed. Results highlight a strong socioeconomic gradient in mortality. High-income countries have an average standardized mortality rate of 1.5 per 100,000 inhabitants, compared to 3.7 per 100,000 inhabitants in lower-middle-income countries. The mortality-to-incidence ratio decreases from 0.45 to 0.15 with increasing income level. Multivariate analyses confirm a significant association between the Human Development Index and cancer mortality among adolescents and young adults (AYAs), after adjusting for other model variables. Joinpoint analyses also reveal heterogeneous temporal trends according to sex and cancer type. This study highlights importance of socio-economic determinants in international inequalities in cancer mortality among AYAs.

## Introduction

Cancer is now one of the leading causes of morbidity and mortality worldwide. According to the most recent estimates from the Global Cancer Observatory, nearly 20 million new cancer cases and approximately 10 million deaths were recorded globally in 2022. Despite progress in prevention, screening, and treatment, the burden of cancer continues to increase due to demographic aging, global population growth, and evolving environmental and behavioral risk factors. While cancers are traditionally considered diseases of the elderly, their impact on adolescents and young adults (AYAs), generally defined as individuals aged 15 to 39, is generating increasing interest, even though the age limits used may vary depending on the country and the databases used. According to [1–3], nearly 1.3 million new cases of cancer and 377,621 deaths were recorded among adolescents and young adults worldwide in 2022. Globally, estimated a standardized incidence rate of 40.3 per 100,000 population and a standardized mortality rate of 11.8 per 100,000 population in this population. Cancers in adolescents and young adults (AYAs) have specific characteristics, that often differ from those of pediatric and adult cancers, with a distinct distribution of tumor sites, different risk factors and the need for tailored care pathways. Leukemias, lymphomas, breast, cervical, thyroid, testicular, and ovarian cancers, as well as sarcomas and certain central nervous system tumors, are among the most frequently observed cancers in this population. Beyond their prevalence, cancers occurring in adolescents and young adults (AYAs) represent a major public health challenge due to their long-term consequences on quality of life, mental health, fertility, professional integration, and social trajectories. This stage of life corresponds to a transitional phase marked by the need for autonomy, the importance of body image, the role of peers, the pursuit of education, entry into the job market, and even the development of life and family plans. Therefore, the onset of cancer during this pivotal period of life has major and lasting repercussions, both at the individual and societal levels, and a lack of awareness or training among primart care providers regarding cancer in adolescents and young adults even in developed contries.

Several recent studies also suggest a worrying trend in the cancer burden among adolescents and young adults (AYAs), which estimates that the global cancer burden among AYAs could increase by approximately 12% between 2022 and 2050, highlighting the growing importance of this population in prevention and management care strategies [2]. Similarly, a study conducted in France reported an increase in the incidence of several cancers in 15- to 39-year-olds between 2000 and 2020, notably colorectal, breast, and thyroid cancers, as well as certain lymphomas and glioblastomas [4]. These trends raise questions about the factors that may explain the increases in incidence and the differences observed between populations and between countries. Socioeconomic factors are one of the hypotheses explaining these disparities. However, in adolescents and young adults, other specific factors may also contribute to the observed differences, including detection methods, the relative rarity of certain cancers, the frequent presence of nonspecific symptoms, diagnostic delays, and a lack of awareness or training among primary care providers regarding cancer in adolescents and young adults, even in developed countries. Since the first works devoted to the topic [6–16], numerous studies populations show that the level of human development, universal health coverage, access to care, healthcare expenditures, and urbanisation can all have an impact at various stages of the care pathway, from prevention to diagnosis, treatment, and post-treatment monitoring.

In the field of cancer, these inequalities often translate into an epidemiological paradox. The most developed countries generally have higher incidence rates, reflecting better case detection and more efficient healthcare systems. Several studies suggest, however, that the increase in certain lifestyle factors, such as obesity, sedentary behavior, or specific environmental exposures, could contribute to the rising incidence of several cancers among young adults in high-income countries. At the same time, resource-limited countries have lower incidence rates but higher mortality rates and less favorable mortality-to-incidence ratios, suggesting delays in diagnosis and challenges related to access to a care of good quality. In [3], it is reported a mortality-to-incidence ratio ranging from 12% in high-income countries to 57% in low-income countries, illustrating the extent of inequalities in care [4]. These studies have notably shown that socioeconomic inequalities remain a major determinant of cancer mortality internationally, however, despite a growing interest in cancers among adolescents and young adults (AYAs), several gaps remain in the literature o this topic. Indeed, the majority of studies focus on a particular tumor site, a limited number of countries, or relatively short study periods. To our knowledge, few studies have simultaneously explored multiple cancer sites, temporal trends, socioeconomic determinants, and sex differences from an international perspective spanning more than two decades. The mechanisms through which socioeconomic determinants influence cancer mortality in AYAs thus remain insufficiently documented.

In this study, our primary objective is to determine whether the level of socioeconomic development is a major factor influencing cancer mortality among adolescents and young adults, independent of any other difference observed between countries. To this end, we analyzed socioeconomic determinants and cancer mortality data for AYAs in 37 countries across all development categories and continents, between 2000 and 2023. Secondary objectives include describing the geographic distribution of the cancer burden among adolescents and young adults, identifying the different socio-epidemiological profiles of countries, examining temporal trends in mortality by sex and cancer type, and assessing the association between various socioeconomic determinants and cancer mortality using panel data models.

## 1. Materials and Methods

### Study Design

This longitudinal ecological study aims to analyze the socioeconomic inequalities associated with cancer mortality among AYAs, as well as their evolution over time between 2000 and 2023. The analyses have been conducted at the population level in several countries, using harmonized international databases.

### Study Population

The study population consists of adolescents and young adults (AYAs), defined as patients aged 15 to 39 years with cancer, according to oncohematology classifications. The study covers 37 countries selected based on the availability and completeness of data for the period 2000–2023, across all development categories and continents. The analyses covered all cancers, excluding non-melanoma skin cancers, for which reporting varies too widely from one country to another and which have a low incidence. Particular attention has been also paid to several specific cancers that are frequently observed in AYAS, including breast, cervical, ovarian, colorectal, testicular, thyroid, and central nervous system cancers, as well as leukemias, lymphomas, and sarcomas.

### Data Sources

Epidemiological data have been extracted from the Global Burden of Disease (GBD) 2023 database, coordinated by the Institute for Health Metrics and Evaluation (IHME), based in Seattle, Washington. Data are available by country, year, sex, cancer cause, and five-year age groups. Socioeconomic indicators have been obtained from the World Bank and from the United Nations Development Programme (UNDP).

### Data Preparation and Harmonization

Data from GBD, World Bank and UNDP were harmonized prior to analysis to ensure consistency in geographic identifiers and the variables used. Since the “Low Income” category is represented by only one country (Togo), it is grouped with the “Lower Middle Income” category to ensure a sufficient sample size for statistical comparisons. However, the results for this group should therefore be interpreted with caution.

### Variables studied

The main indicators studied are:

- Age-standardized mortality rates (ASMR);

- Age-standardized incidence rates (ASIR);

- Mortality-to-incidence ratio (MIR).

The main analyses focused on ASMR. The socioeconomic variables studied included:

- Human Development Index (HDI);

- Percentage of public health expenditure in global health expenditure;

- Global health expenditure (% of GDP);

- Level of urbanisation;

- Universal Health Coverage (UHC) index.

### Construction of indicators

The age-standardized rate of an observable variable *X* like the mortality or the incidence of a disease (*ASXR*) is calculated by direct standardization from age-specific rates. Standardization has been performed by applying the weights of the WHO standard reference population to the different age groups between 15 and 39 years, according to the following formula:

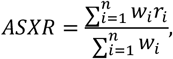

where *ri* represents the specific rate of the observable *X* for the class *i* and *wi* the weight of the class *i* in the standard population. Then, the ratio mortality *M* / incidence *I* (*MIR*) is calculated following the formula:

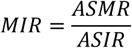

### Statistical methodology

A descriptive analysis of epidemiological and socioeconomic indicators has been first conducted using maps, comparative graphs, and time-series visualizations. Descriptive analyses, maps, correlations, principal component analysis (PCA), and unsupervised classification methods (in which the classes are unknown a priori and are identified solely using the raw data, then interpreted using metadata external to the classification) were performed using data from the most recent year of observation (2023), in order to characterize the recent socio-epidemiological profiles of the countries studied.

Exploratory multivariate analyses were then conducted using:

- principal component analysis (PCA);

- and unsupervised classification (k-means clustering).

K-means clustering has been performed on the factor coordinates derived from PCA. The optimal number of clusters has been explored using the elbow method and the silhouette score. Hierarchical clustering based on principal components (HCPC) then used to visualize and confirm the structure of the resulting groups.

The differences in ASMR observed between clusters were assessed using Welch’s ANOVA, which is more appropriate when variances are heterogeneous. Tukey’s post-hoc comparisons were then performed.

Temporal trends were studied using:

- Estimated Average Percentage Change (EAPC);

- and Joinpoint analyses used to estimate Annual Percentage Changes (APC) and Average Annual Percentage Changes (AAPC).

Fixed-effects and random-effects panel models were also estimated to examine the associations between socioeconomic variables and cancer mortality among AYAs.

### Used softwares

Statistical and graphical analyses have been performed using R (version 4.5.2), primarily using the following packages: tidyverse, ggplot2, fixest, plm, FactoMineR and factoextra. Joinpoint analyses have been performed using the Joinpoint Program software developed by the National Cancer Institute (NCI, USA).

## Results and interpretation

### Socioeconomic level

Significant disparities in incidence and mortality indicators are observed among the countries studied and according to their level of socioeconomic development.

Table 1 shows that urbanisation is proportional to cancer incidence, and we also know that urbanisation is linked to high noise levels, exposure to air pollution, a diet high in ultra-processed foods, a sedentary lifestyle, and social isolation—all of which are major factors contributing to comprehensive morbidity and mortality. The descriptive characteristics of the 37 countries in 2023 highlight a socioeconomic gradient correlated with the cancer burden among AYAs. High-income countries have an average age-standardized incidence rate (ASIR) of 11.4 per 100,000 population, compared with 9.7 in lower-middle-income countries. Conversely, the age-standardized mortality rate (ASMR) decreases sharply with income level, falling from 3.7 per 100,000 population in lower-middle-income countries to 1.5 in high-income countries. The mortality-to-incidence ratio (MIR) follows the same trend, with an average value of 0.45 in lower-middle-income countries compared with only 0.15 in high-income countries. The most affluent countries also have higher Human Development Index (HDI) levels (0.93 versus 0.62), health coverage (86.8 versus 55.7), and health expenditure (10.3% versus 4.4% of GDP).

**Table 1.** Characteristics of 37 countries depending on their income group in 2023.

| Variable | Lower Income | Middle Income | High income |
|---|---|---|---|
| ASIR | 9.7 ± 3.2 | 9.6 ± 2.5 | 11.4 ± 2.6 |
| ASMR | 3.7 ± 1.3 | 2.9 ± 0.8 | 1.5 ± 0.3 |
| MIR | 0.45 ± 0.06 | 0.33 ± 0.09 | 0.15 ± 0.05 |
| HDI | 0.62 ± 0.08 | 0.79 ± 0.04 | 0.93 ± 0.04 |
| Urbanisation rate | 47.2 ± 11.5 | 75.0 ± 12.4 | 81.9 ± 8.4 |
| GDP % Health expenditures | 4.4 ± 1.2 | 6.3 ± 2.4 | 10.3 ± 2.2 |
| Public expenditure | 33.6 ± 14 | 60.4 ± 12 | 69.9 ± 14 |
| UHC index | 55.7 ± 10 | 76.1 ± 8 | 85.8 ± 3.1 |

**Table 2.** Random-effects panel model with interaction between the HDI and gender.

| Variable | Coefficient | SE robuste | Statistique t-test | p-value |
|---|---|---|---|---|
| Constant | 4.231 | 0.333 | 12.890 | < 0.001 |
| HFI Women | -1.462 | 0.650 | -2.249 | 0.025 |
| HDI Man | -1.700 | 0.225 | -7.571 | < 0.001 |
| Public expenditures | 0.002 | 0.003 | 0.768 | 0.431 |
| Urbanisation | -0.009 | 0.005 | -1.741 | 0.082 |
| Interaction HDI/Gender | 1 .876 | 0.253 | 7.420 | < 0.001 |

Random effects: robust clustered standard errors at the country level. Dependent variable: log(ASMR). Hausman test: p = 0.203; Wald test for the interaction: p < 0.001.

### Geographic Distribution

The geographic distribution of incidence and mortality indicators reveals significant disparities among the countries studied (Figure 1).

**Figure 1.**
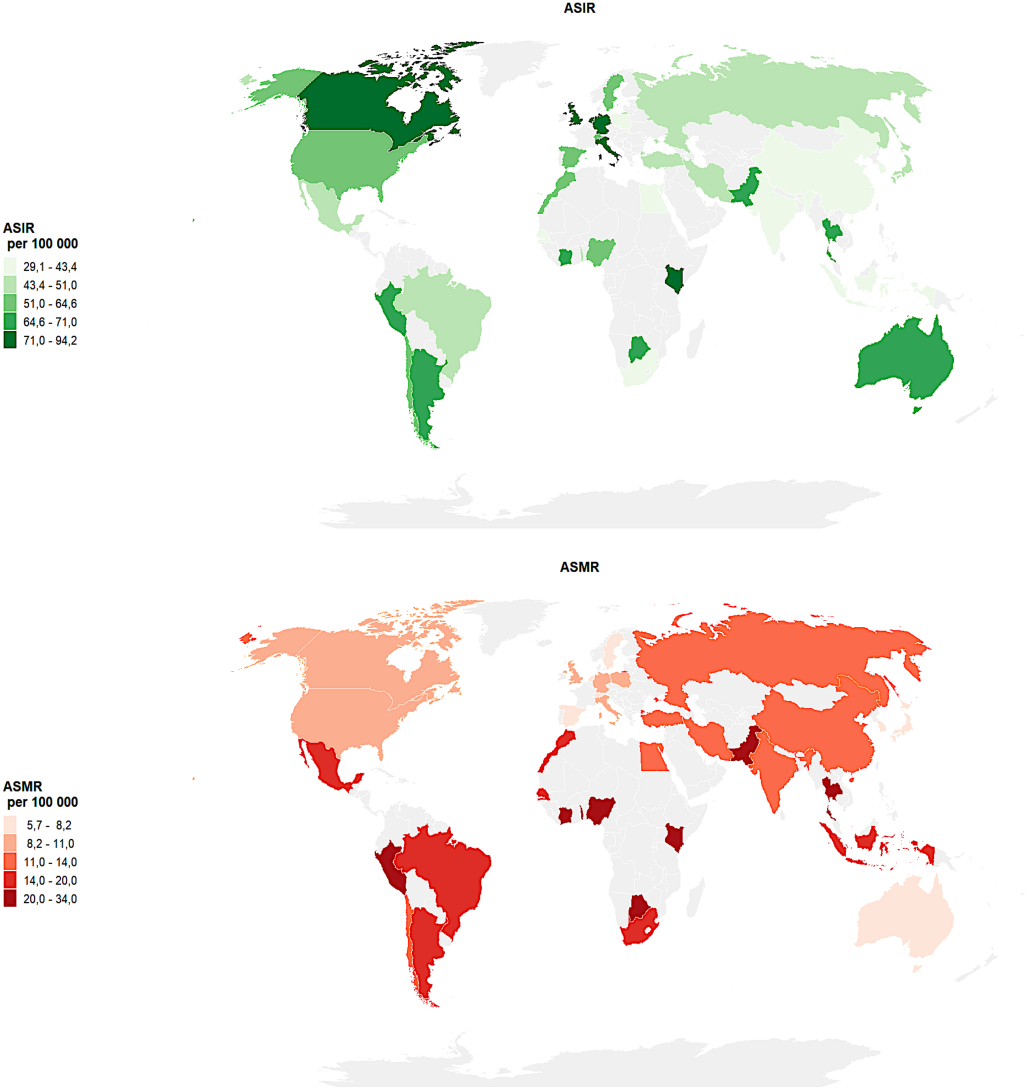
Geographic distribution of ASIR and ASMR among AYA in 2023.

The highest ASIR rates are primarily observed in several high-income countries, particularly in North America, Western Europe, and Oceania. Conversely, the highest ASMR rates are found primarily in several countries in Latin America, sub-Saharan Africa, and Southeast Asia [9]. This geographic distribution highlights significant international heterogeneity in the cancer burden among AYAs, suggesting the existence of substantial differences between health care systems, their diagnostic capabilities, and treatment conditions across different regions of the world. The profiles of the most common cancers also varied by region of the world and by gender. In terms of incidence, breast and cervical cancers account for a significant portion of the burden observed in several countries like Brazil, China, India and Russia, while leukemias and central nervous system cancers are among the leading causes of mortality in many other countries studied like Australia, Spain, UK and USA. Testicular cancer also hasa much higher incidence in several high-income countries. Detailed maps of the most common cancers by incidence and mortality are presented in the Appendix.

### Incidence-Mortality Paradox

The mortality-to-incidence ratio (MIR), commonly used as an indirect indicator of prognosis and the overall performance of cancer care systems, decreases significantly (p < 0.001) with increasing human development (Figure 2). Countries with low or medium HDI values have median MIR values close to 0.6, compared with approximately 0.1 to 0.2 in countries with very high HDI values, suggesting a generally less favorable prognosis for cancer among AYAs in the context of less developed countries. These disparities are particularly concerning in this population, for whom delays in diagnosis and access to specialized care pathways remain significant challenges.

**Figure 2.**
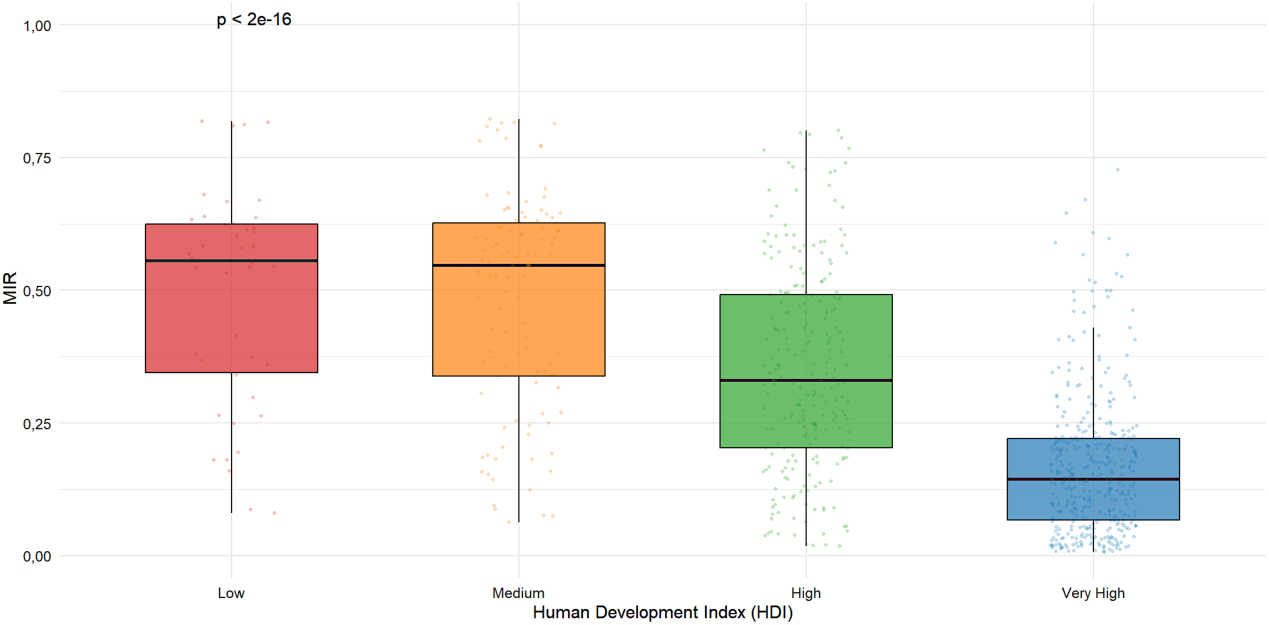
Socioeconomic gradient of MIR for AYAs (2023).

In Figure 2, the minimal difference between low-and middle-income countries can be explained by the small number of low-income countries.

### Correlations

To explore the relationships between socioeconomic and health indicators, a Spearman’s correlation analysis has been conducted between the main variables studied (Figure 3).

**Figure 3.**
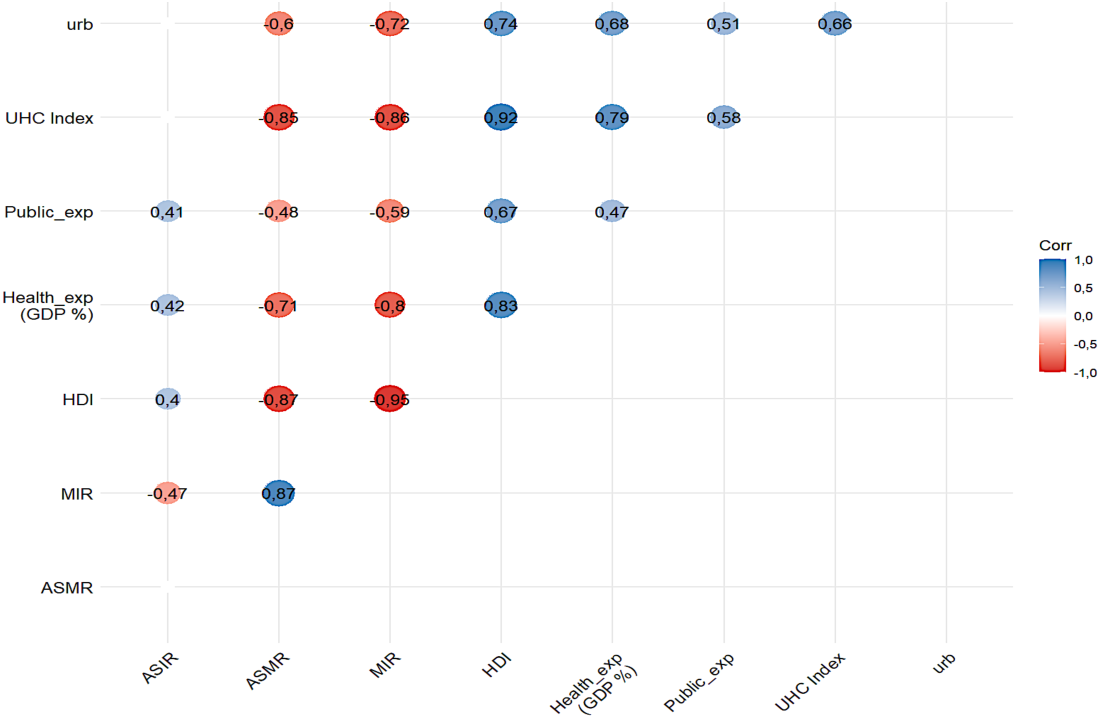
Spearman’s correlations.

**Figure 4.**
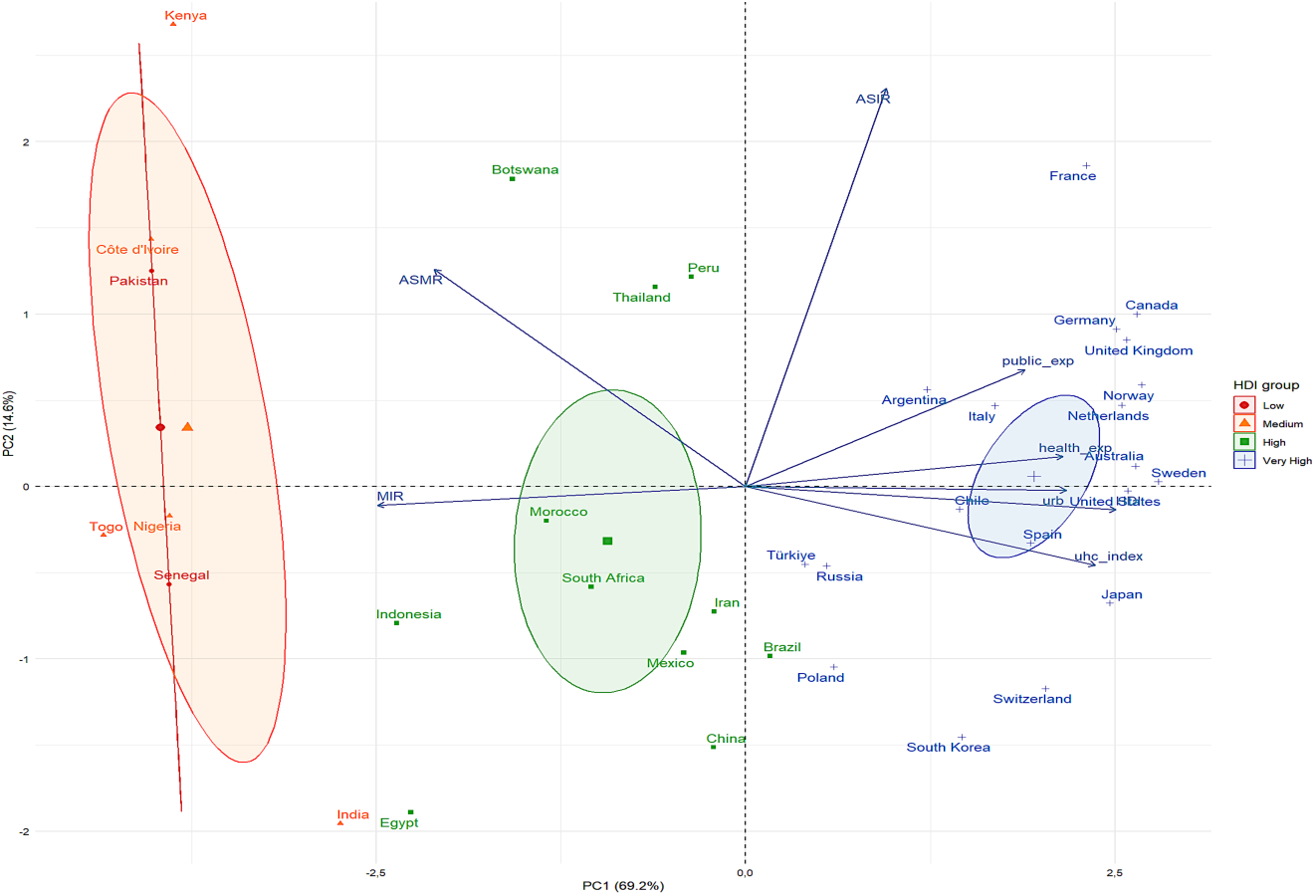
Representation of the variables for the 37 studied countries on the two first principal components.

Spearman’s correlation analyses reveal strong associations between socioeconomic indicators and the cancer burden among AYAs (Figure 3). The MIR shows the strongest negative correlations with the Human Development Index (r = -0.72), the Universal Health Coverage Index (r = -0.86), health expenditure as a percentage of GDP (r = -0.80), and the level of urbanisation (r = -0.72). Similarly, the standardized mortality rate is strongly and negatively correlated with the HDI (r = -0.87), UHC (r = -0.85), and health expenditure (r = -0.71). The standardized incidence shows moderate positive correlations with key indicators of socioeconomic development, notably the HDI (r = 0.40), health expenditure (r = 0.42), and the share of public health expenditure (r = 0.41). Finally, the MIR is strongly correlated with the ASMR (r = 0.87), suggesting that the differences observed between countries in terms of relative mortality may reflect variations in survival, access to diagnosis, quality of care, and the organization of care pathways among AYAs.

### Principal Component Analysis and Unsupervised Classification

Principal Component Analysis (PCA) To explore the overall structure of countries based on their epidemiological and socioeconomic characteristics, a principal component analysis (PCA), followed by an unsupervised classification, has been performed. The first two principal components alone account for 83.8% of the total variance in the data (69.2% for PC1 and 14.6% for PC2), indicating a satisfactory representation of the data structure. The first component contrasts socio-health development indicators (HDI, UHC Index, health expenditure, and urbanisation) against relative mortality indicators (ASMR and MIR). This axis can be interpreted as a gradient of socio-health development, contrasting countries with more developed health systems and better access to cancer care with countries facing higher relative mortality among AYAs. The second component shows more differences between incidence and mortality, potentially reflecting variations in the epidemiological profiles of the studied countries.

The plotting of countries on the first factor graph reveals a distinct trend based on their level of socioeconomic development. Countries with low and medium HDI scores are predominantly located in the negative portion of Axis 1, which is associated with higher levels of ASMR and MIR. Countries with high or very high HDI are mainly located in the positive part of this axis, characterized by higher levels of universal health coverage, health spending, and urbanisation. This pattern is particularly evident for countries with very high HDI, such as France, Canada, Norway, and the United Kingdom, which form a relatively homogeneous group on the factor plot. In contrast, several less-developed countries, notably Kenya, Côte d’Ivoire, Pakistan, and Senegal, are distinguished by profiles associated with higher relative mortality.

The second factor axis explains a smaller proportion of the variance (14.6%) and appears to distinguish countries that sometimes have comparable levels of development but different epidemiological profiles. This component may reflect differences in the distribution of the most common cancers among AYAs across countries.

Unsupervised classification To identify homogeneous country profiles, a hierarchical principal component classification (HCPC) has been performed using the factor coordinates derived from PCA. This analysis identified three clusters with distinct socio-epidemiological characteristics (Figure 5).

**Figure 5.**
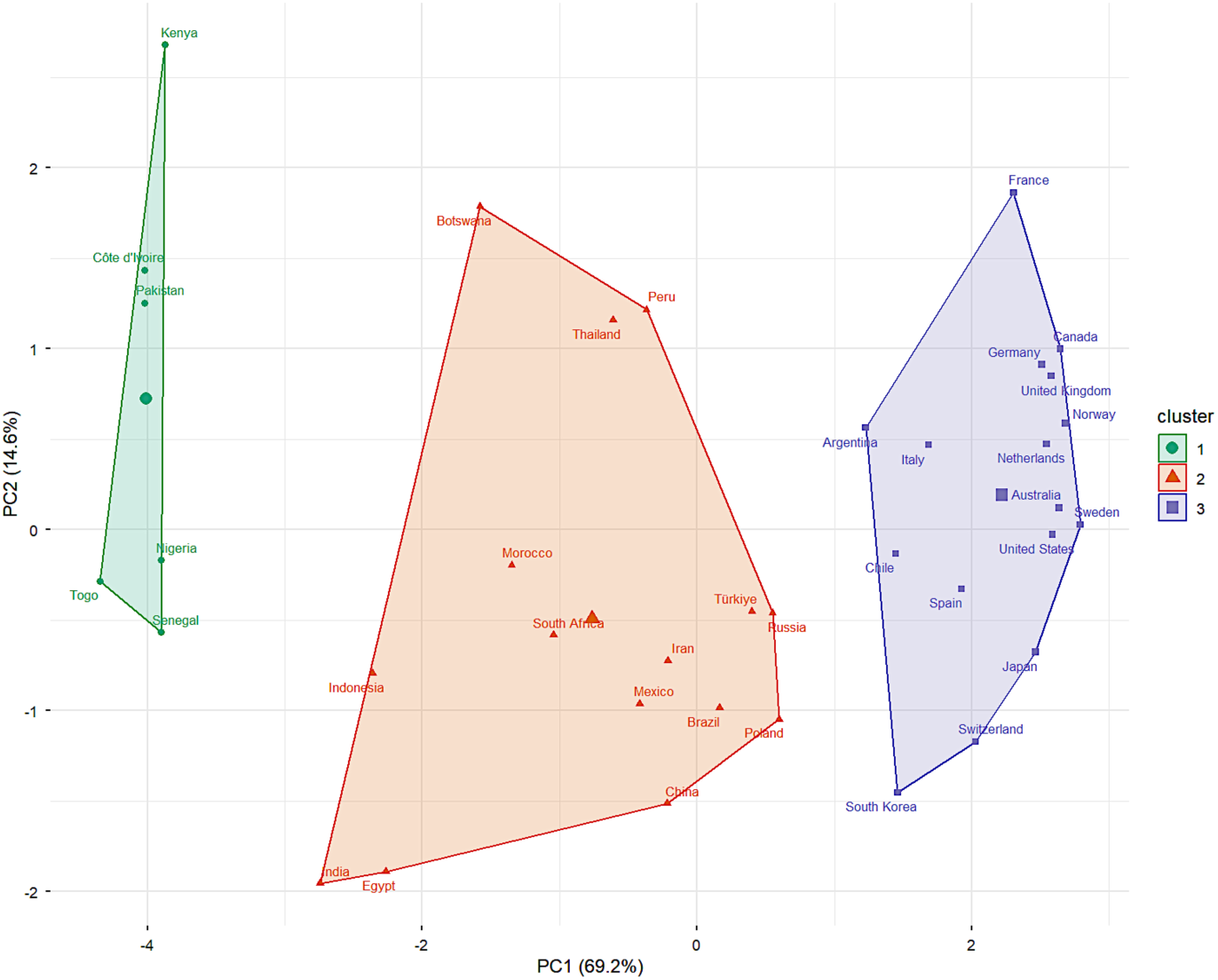
Hierarchical classification on principal compnents for the 37 studied countries.

Cluster 1 comprises six countries primarily characterized by a low level of socio-health development. This group has the highest levels of age-standardized mortality (ASMR = 24.8 per 100,000 population) and mortality-to-incidence ratio (MIR = 0.51), combined with the lowest values for the HDI (0.569), universal health coverage (49.3), and health expenditure (4.2% of GDP). These characteristics may reflect a less favorable environment in terms of prevention, information and specific training explaining early diagnosis and appropriate therapeutic management of cancer among adolescents and young adults with cancer as well as more general differences in the organization and resources available to health care systems.

Cluster 2, comprising fifteen countries, occupies an intermediate position with an average ASMR of 15.5 per 100,000 population and an average MIR of 0.36. The socioeconomic indicators are also intermediate (average HDI = 0.780; UHC = 75.0; health expenditure = 5.9% of GDP), suggesting more heterogeneous socio-epidemiological transitions with contrasting profiles of the burden of cancer among AYAs.

Cluster 3 comprises sixteen countries with the most favorable profiles. It is characterized by the highest levels of HDI (0.934), universal health coverage (86.0), health expenditure (10.8% of GDP), and urbanisation (84.4%), combined with the lowest levels of mortality (ASMR = 8.95 per 100,000 inhabitants) and relative mortality (MIR = 0.14). The identified groups reveal a consistent gradient of socio-health development, ranging from less advantaged countries to those with the most effective health systems. The differences in ASMR observed among the three clusters are statistically significant (Welch’s ANOVA, p < 0.001), and post-hoc comparisons confirm significant differences among each of the three clusters. However, the clusters obtained do not perfectly align with the economic classifications typically used. Some countries with comparable income levels belong to different groups (like Turkey and Russia in group 2), suggesting that inequalities in cancer mortality among young and middle-aged adults do not depend solely on the level of national wealth, but also on more complex health and organizational factors, such as access to specialized care or the organization of care pathways.

### Temporal Trends

Temporal trends in cancer mortality among AYA were then examined over the 2000–2023 period using log-linear models to estimate the Estimated Annual Percent Changes (EAPC) from the age-adjusted cancer mortality rates. The results highlight heterogeneous trends depending on cancer type, gender and human development level.

On Figure 6, the dots represent the estimated EAPCs, and the horizontal bars represent their 95% confidence intervals. A negative value indicates an average decrease in mortality over the study period. For all cancers combined, countries with a very high HDI show the most favorable trends, with an average decrease in mortality of 1.32% per year (EAPC = -1.32%). This decline is more moderate in countries with medium (–0.36%) and high (–0.26%) HDI, while countries with low HDI show an overall stable trend (EAPC = +0.08%). These results suggest faster progress in controlling cancer mortality in the most favorable socio-health contexts. Trends also vary by cancer site. Hematologic cancers show the most favorable trends, particularly in the countries with very high HDI. Leukemias show an average annual decrease of 2.05%, while Hodgkin’s lymphomas show a decline of nearly 3% per year (EAPC = -2.96%). These results may reflect the therapeutic advances observed over the past few decades for these diseases, particularly thanks to improved treatment protocols and better structured care for AYAs in certain countries. Conversely, several types of cancer show less favorable trends in low- and middle-income countries. Breast cancer, in particular, shows an average increase in mortality in countries with a low HDI (EAPC = +1.40%) and a medium HDI (+1.02%). Similar trends are observed for ovarian cancer, with estimated EAPCs of 1.18% and +1.12 %, respectively. However, the causes are likely different: since ovarian cancer is often diagnosed at an advanced stage, so rather than the location of the tumor, it is likely the even longer delays in diagnosis that explain the excess mortality. Overall, the trends observed differed by sex and cancer type, reflecting contrasting situations among the countries studied.

**Figure 6.**
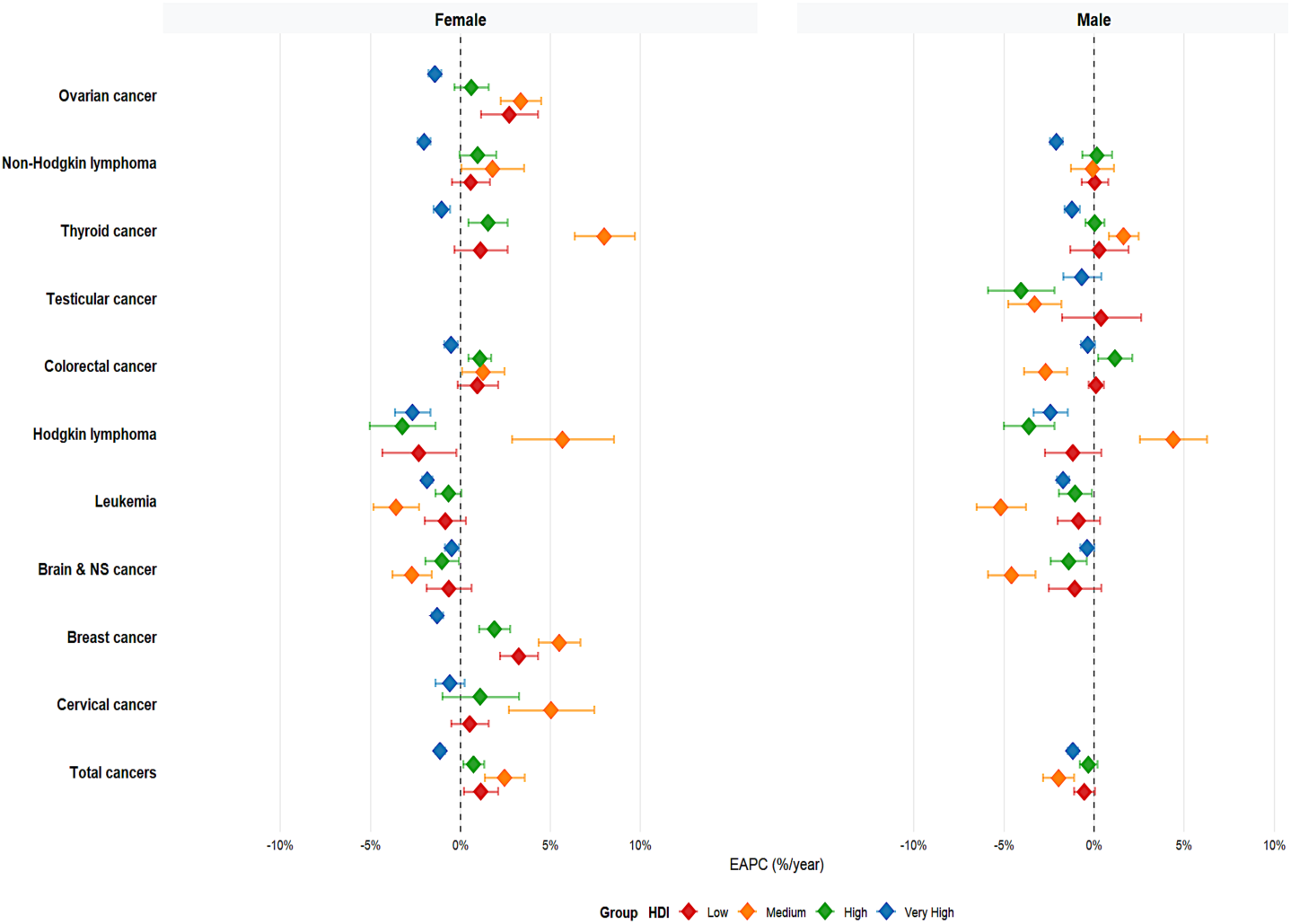
Trends in cancer mortality among AYAs by HDI and cancer site (EAPC).

To explore possible breaks in trends during the study period, a analysis has been conducted on age-standardized mortality rates. This approach makes it possible to identify changes in the temporal evolution of trends and to estimate annual percentage changes (APC) by segment as well as overall average trends (AAPC). The analyses reveal heterogeneous temporal dynamics depending on the type of cancer and gender. Some cancers exhibit stable trends during the study period, while others show breaks in trends.

Among women, cervical cancer is a particularly telling example of these nonlinear trends (Figure 7A). Despite an overall decline in mortality over the entire period (AAPC = -0.46%/year; 95% CI [-0.82; -0.21]), they reveal three distinct phases: a marked decline between 2000 and 2007 (APC = −4.52%/year; 95% CI [−6.46; −3.52]), followed by a period of stabilisation between 2007 and 2020 (APC = 0.79%/year; 95% CI [−2.20; 1.27]), and then a recent increase after 2020 (APC = 3.95%/year; 95% CI [1.12; 5.77]). Among men, colorectal cancer also shows several changes in trend over the study period (Figure 7B). While mortality decreases significantly between 2000 and 2009 (APC = −1.57%/year; 95% CI [−2.16; -1.25]), a stabilization phase is observed between 2009 and 2019 (APC = 0.42%/year; 95% CI [-0.38; 0.79]), followed by a recent increase between 2019 and 2023 (APC = 2.41%/year; 95% CI [1.32; 3.53]). Over the entire period, however, the average trend remains stable (AAPC = -0.02%/year; 95% CI [-0.16; 0.08]). Overall, the results of the analyses remain consistent with the average trends observed previously. However, this approach makes it possible to highlight more nuanced changes over time, with some locations showing less favorable recent phases despite an overall decline observed throughout the entire period. The detailed results of the APC, AAPC, and their 95% confidence intervals are presented in the Appendix.

**Figure 7A.**
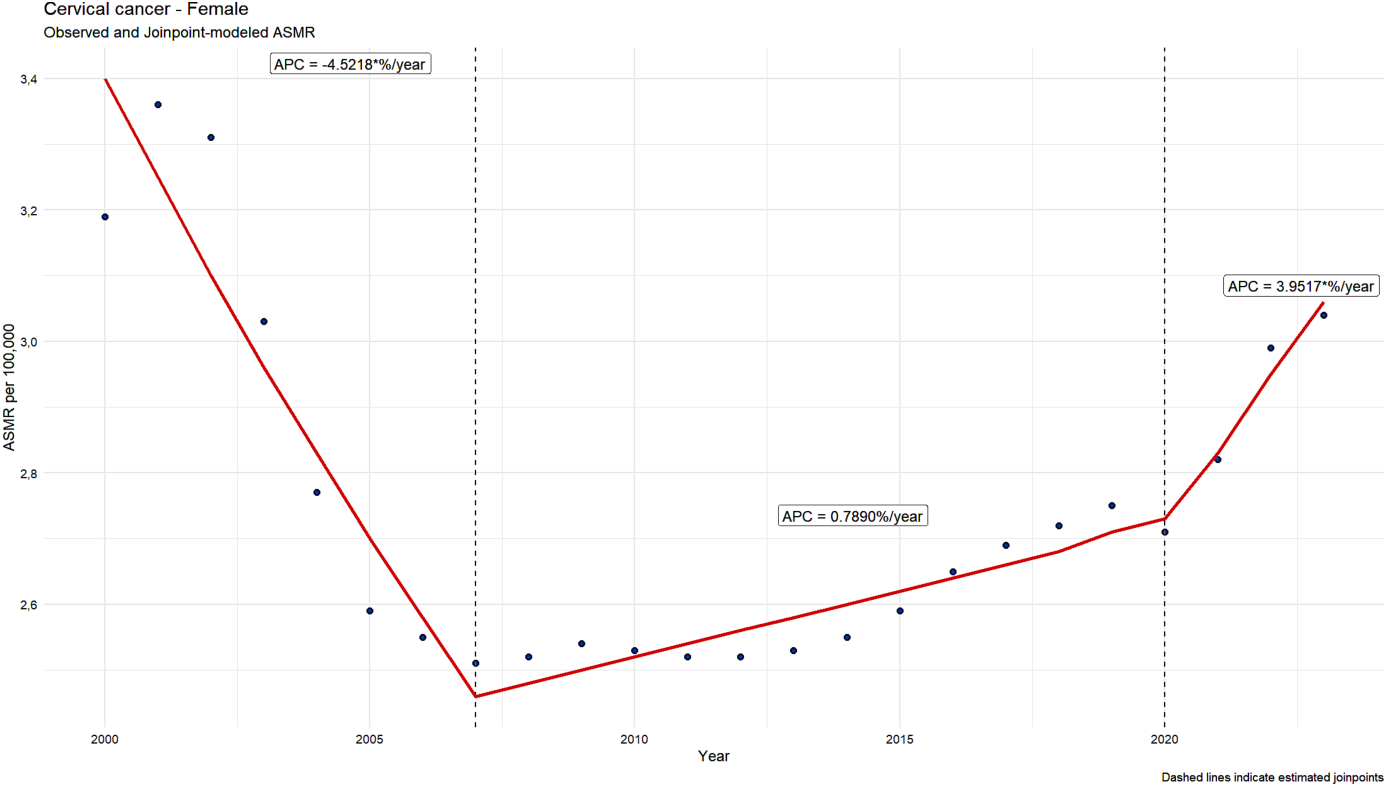
Trends in age-standardized mortality rates and trend changes identified by Joinpoint analysis among AYAs for cervical cancer in females.

**Figure 7B.**
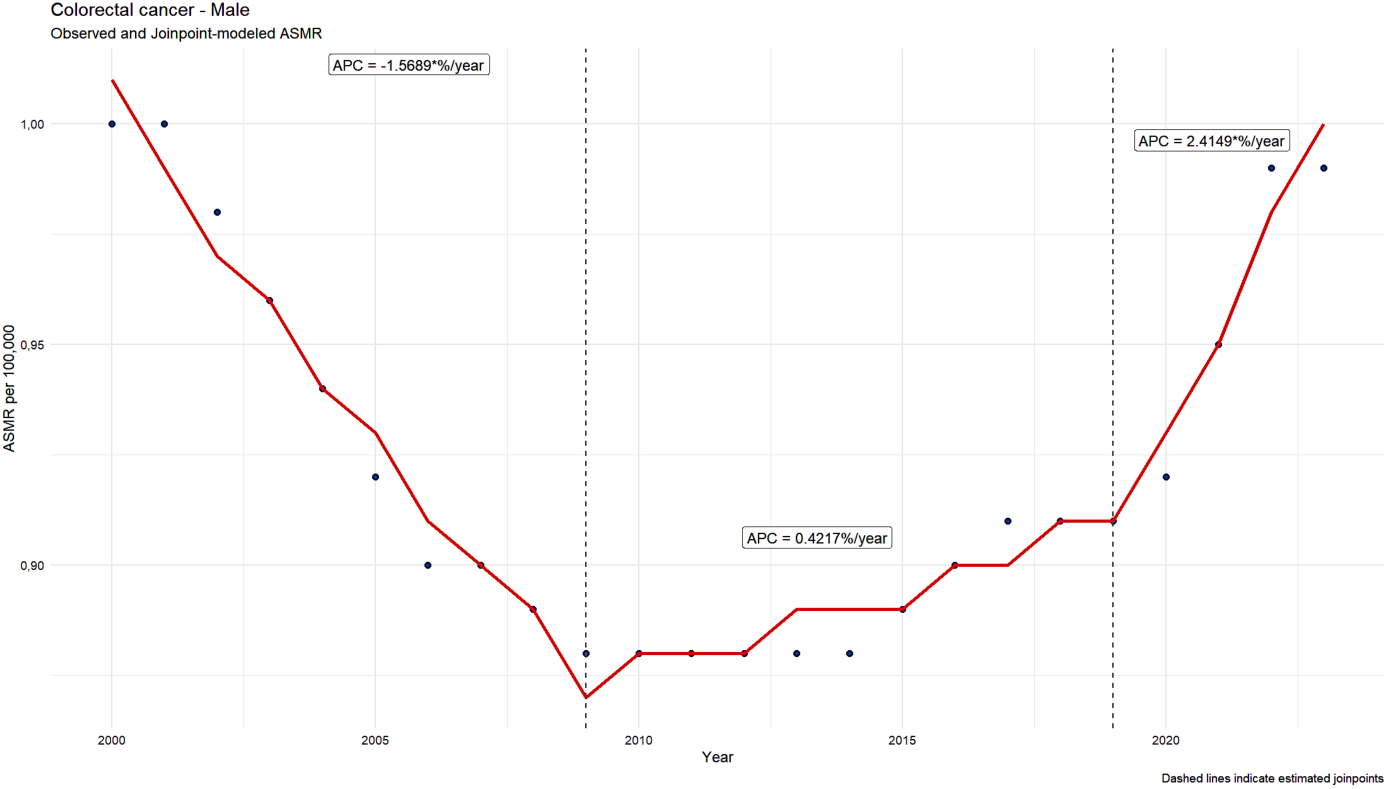
Trends in age-standardized mortality rates and trend changes identified by Joinpoint analysis among AYAs for colorectal cancer in males.

**Figure 7C.**
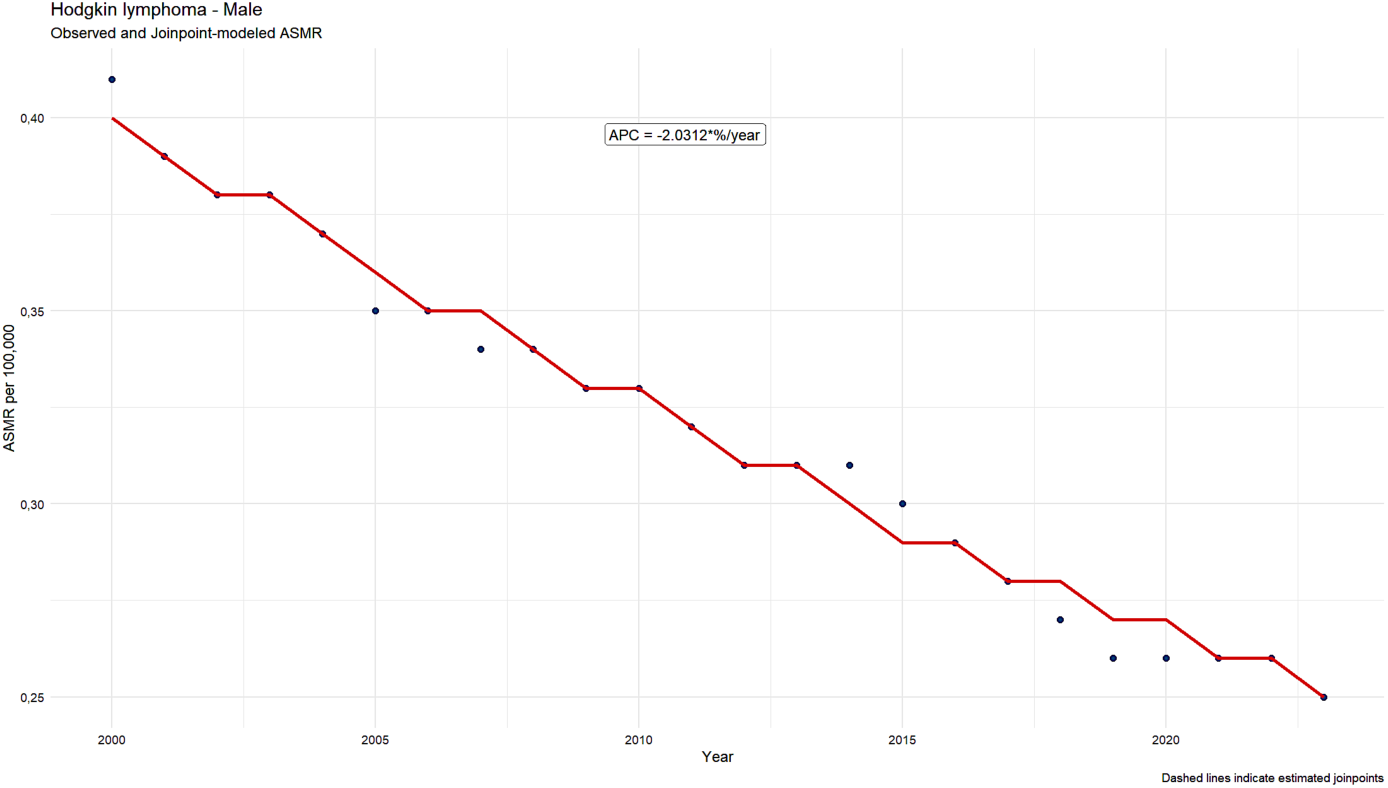
Trends in age-standardized mortality rates and trend changes identified by Joinpoint analysis among AYAs for Hodgkin lymphoma in males.

### Panel Models

To explore the association between socioeconomic factors and cancer mortality among AYAs, regression models using panel data were then conducted. These analyses make it possible to simultaneously account for the temporal dimension (2000–2023) and the differences observed across countries. The models include, in particular, the Human Development Index (HDI), health care spending (% of GDP), the share of public health care spending, gender, the urbanisation rate, and the Universal Health Coverage (UHC) index as explanatory variables for standardized mortality rates. Due to a strong correlation with the share of public health expenditure, total health expenditures (% of GDP), as well as the universal health coverage index—which is strongly correlated with the HDI—were excluded from the main models to limit multicollinearity issues (that is, with increased influence on the joint variable from highly correlated variables).

Fixed-effects and random-effects models were estimated and then compared using the Hausman test. Given the results of the Hausman test (p = 0.203), random-effects models were preferred for the main analyses. Fixed-effects models were nevertheless retained as part of sensitivity analyses to assess the robustness of the results obtained. To account for heteroscedasticity and intra-country correlations over time, standard errors were corrected using robust estimators clustered at the country level (that is, largely independent of the nature of the empirical distributions and the sample sizes). An interaction between gender and the HDI was then explored to assess whether the association between socioeconomic development and mortality varied by gender (due to sociological differences, for example a women’s poorer access to healthcare). Adding the interaction term significantly improved the model (Wald test, p < 0.001).

Random-effects panel models show a significant association between the level of socioeconomic development and cancer mortality among AYAs. Among women, an increase in the HDI is associated with a significant decrease in standardized mortality (β = -1.462; p < 0.025), a trend also observed among men (β = -1.700; p < 0.001). The interaction between the HDI and gender is also significant (β = 1.876; p < 0.001), indicating that the observed association between socioeconomic development and mortality differs by gender and appears less pronounced among men or means that the impact of socioeconomic status on cancer mortality among AYAs is greater among women. The share of public health expenditure is not significantly associated with mortality after adjusting for the other variables in the model (β = 0.002; p = 0.431). Similarly, urbanization does not show a statistically significant association, although a negative trend was observed (β = -0.009; p = 0.082).

Overall, human development is the socioeconomic variable most strongly associated with cancer mortality among AYA in the countries studied. However, the observed associations differ by gender, suggesting that the mechanisms linking socioeconomic conditions to mortality may vary between women and men.

## Discussion

The objective of this study was to explore the socioeconomic factors associated with cancer mortality among adolescents and young adults (AYAs) in 37 countries, based on data available from 2000 to 2023. The results highlight significant international disparities, characterized by a marked socioeconomic gradient that significantly impacts cancer mortality among 15-to 39-year-olds. The data underlying these results come from official databases; however, it is not always possible to guarantee their completeness, consistency, and freedom from major biases, which may partly explain these disparities. It should be noted, for example, that the 15–39 age group is heterogeneous from an epidemiological and therapeutic standpoint: when breaking down the group into those aged 15–24 and those aged 25–39, significantly different results are observed in terms of tumor location, care pathways, and mortality. Countries with the highest levels of human development, universal health coverage, and health care spending generally have the lowest levels of age-standardized mortality and mortality-to-incidence ratios.

Correlation, classification, and panel analyses all point to a robust association between socioeconomic development and cancer mortality among AYAs. The correlations observed between the HDI and the standardized mortality rate (r = -0.87), as well as between UHC and mortality (r = -0.85), illustrate the strength of this relationship. These results appear consistent with the conceptual framework of the social determinants of health [5], according to which social, economic, and health conditions profoundly influence the health status of populations.

The HDI, which simultaneously incorporates dimensions related to health, education, and standard of living, can be considered a composite indicator of these structural determinants. In our study, the strong correlations observed between the HDI and mortality indicators, as well as the results of the panel models, suggest that these mechanisms also play an important role in the inequalities in cancer mortality observed among AYAs. These mechanisms could notably influence diagnostic delays, access to specialized centers, clinical trials, and innovative treatments. The differences observed according to the level of human development suggest the existence of major inequalities in cancer care among AYAs. In our study, countries in the least advantaged cluster had an average standardized mortality rate of 24.8 per 100,000 population, compared with 8.95 per 100,000 population in the most advantaged cluster—a mortality rate nearly three times higher.

Similarly, the mortality-to-incidence ratio ranged from 0.511 to 0.14 between these two groups. These results suggest that the differences observed between countries do not solely reflect variations in cancer incidence, but also inequalities in access to diagnosis, specialized treatments, and continuity of care. This finding is consistent with previous studies, which have shown that socioeconomic inequalities remain a major determinant of cancer mortality on an international scale [4]. Our analyses are also consistent with certain other studies from the literature about AYAs cancers, which have reported marked differences in mortality between countries based on their income levels, with a mortality-to-incidence

In France, a study [2] reported an increase in the incidence of several types of cancer between 2000 and 2020 among 15-to 39-year-olds, particularly colorectal, breast, and thyroid cancers. Similar trends have been described in several high-income countries, suggesting a gradual shift in the epidemiological profile of cancers among AYAs. These trends are particularly concerning among AYAs, given the significant number of potential years of life lost and the long-term consequences of treatments on quality of life, fertility, and social and occupational integration. In our study, certain cancer sites also showed less favorable trends over time, particularly colorectal cancer in men and certain cancers in women (namely thyroid and breast cancers). Several hypotheses have been proposed to explain these trends, including changes in dietary habits, rising rates of obesity and physical inactivity, as well as improvements in diagnostic practices for certain cancer sites, such as thyroid cancer.

The analysis of temporal trends also revealed contrasting patterns across different groups of countries. The most significant declines in mortality were observed in countries with a very high HDI, with an average decrease of 1.32% per year for all cancers, compared with a nearly stable trend in countries with a low HDI (+0.08% per year). At the same time, certain cancer sites showed less favorable trends in countries with low or medium levels of development. Joinpoint analyses also highlighted the nonlinear nature of certain mortality trajectories during the study period [6]. The trend breaks observed for cervical cancer in women and colorectal cancer in men illustrate the complexity of temporal changes in the cancer burden among AYAs. These results fit into an international context marked by a shifting epidemiological profile of cancers among AYAs. In particular, an increase in the incidence of several cancer sites in this population has been reported worldwide [1], while some studies project an overall increase in cancer burden among AYAs of approximately 12% by 2050 [2].

The trends observed in our study may reflect both the progress made in certain countries in terms of prevention and care, as well as the emergence of new epidemiological patterns for certain types of cancer. In particular, some recent changes may have been influenced by disruptions caused by the COVID-19 pandemic, which may have affected access to screening, diagnosis, and cancer care in several countries. However, these trends should be interpreted with caution, given the still-limited historical data available for the period after 2020.

Panel models also confirmed the importance of socioeconomic development in the disparities observed across countries. After adjusting for the other variables in the model, the HDI remained significantly associated with cancer mortality among AYAs. This result suggests that the observed association is not limited to a simple effect of national wealth but likely reflects the combined influence of several dimensions of human development, including health status, educational attainment, and living conditions. The absence of a significant association with public health spending after adjustment could be explained by the comprehensive nature of the HDI, which simultaneously captures several structural determinants of health. Thus, beyond the volume of resources invested in healthcare systems, the results highlight the importance of general socioeconomic conditions in the inequalities observed among AYA [7]. A significant interaction between gender and the HDI also suggested that this association differed by sex. Several mechanisms could explain these differences, including the distribution of cancer sites, health behaviors, patterns of healthcare utilization, and diagnostic pathways. However, the aggregated data used in this study do not allow for a direct exploration of these hypotheses, and further research would be need.

This population also has several distinctive characteristics that may contribute to the observed inequalities. Adolescents and young adults occupy a unique position between pediatric and adult oncology health care. Several studies have highlighted more frequent delays in diagnosis, reduced access to clinical trials, and care that is sometimes less structured than in other age groups. These challenges may contribute to certain disparities observed between countries and between health care systems.

The findings also have several implications for public health. Although France is among the countries with a very high level of human development, certain trends observed call for vigilance. Recent trends identified for testicular cancer in men, as well as changes observed for cervical and breast cancer in women, underscore the importance of continuing prevention and early diagnosis efforts among young adults and adolescents. These observations are consistent with certain studies [3], which have highlighted a recent increase in the incidence of several cancers among young adults and adolescents, particularly colorectal, breast, and thyroid cancers. They also align with the conclusions of the EPI-AJA program, which emphasize the importance of specific monitoring of emerging cancers among young adults [4]. In this context, raising awareness of modifiable risk factors, and improving access to a better access of the youngs to the GP and other health care providers, could help reduce the future burden of cancer in this population. These findings serve as a reminder that, even in countries with well-functioning health care systems, prevention and early detection remain essential strategies for reducing health inequalities and improving cancer outcomes among AYA.

This study has several strengths. It is based on an international approach covering more than twenty years of data and incorporating a variety of socioeconomic contexts. The combined use of descriptive analyses, temporal trend analyses, classification methods, and panel models has made it possible to explore mortality inequalities across several complementary dimensions. To our knowledge, few studies have simultaneously combined international descriptive analyses, PCA, unsupervised classification, Joinpoint analyses, and panel models to examine cancer mortality inequalities among AYAs over a period of more than twenty years. Furthermore, data from the Global Burden of Disease [9] provide the opportunity to compare standardized indicators across countries over a long period.

However, certain limitations must be taken into account. First, this study is based on data aggregated at the country level and therefore does not allow for the establishment of causal relationships at the individual level. The observed associations thus stem from an ecological approach. Furthermore, certain potentially important determinants—such as individual behaviors, environmental exposures, or diagnostic delays—were not available in the databases used. In addition, the Global Burden of Disease [9] estimates are based in part on statistical models whose accuracy may vary depending on the availability and quality of national data. Finally, some cancers had small sample sizes, which may limit the stability of certain temporal estimates.

This study, which allows us to draw comparisons and useful insights based on population data among countries and their health care systems, also highlights the need to take into account other factors that are just as critical to the outcomes of specific patients—especially those with rare diseases such as AYAs:

i. the sincere and active commitment of all stakeholders (patient and professional associations, learned societies, health authorities, and elected officials, among others) to improving care for AYA patients with cancer, as was exemplarily demonstrated in France with the first National Cancer Plans (2003–2007 and 2009– 2014), which led to real and significant progress
ii. changes in regulations and the allocation of financial resources to implement organizational structures that genuinely promote access to and the quality of care for AYAs, in France, for example, with Circular DHOS/SDO/2005/101 of February 22, 2005, on general quality criteria for cancer care, or Instruction DGOS/R3/INCA/ 2016/177 of May 30, 2016, which is more specific to adolescents and young adults with cancer [17]
iii. raising awareness and providing training for frontline healthcare professionals,
iv. taking care not to allow the plight of AYAs to become a marketing opportunity. The NGO « Médecins du Monde », like other patient advocacy groups around the world, regularly denounces in its reports and white papers the unjustified skyrocketing of prices for cancer drugs and therapeutic innovations, which severely limits patients’ access to the care they need and even jeopardizes existing solidarity-based healthcare systems in developed countries. (https://www.medecinsdumonde.org/campaign/le-prix-de-la-vie/).

Healthcare professionals working with AYAs are also very aware of these risks and discussed them extensively at the most recent international conference dedicated to adolescents and young adults with cancer, held in Melbourne in December 2024, with the main theme being : « Ethics, Equity, and Emerging Ideas » (https://gems.eventsair.com/ayaga2024/). The plenary session on the ethics of innovation highlighted the extensive research on the unethical nature of certain industry-sponsored clinical trials, which divert substantial resources in the name of therapeutic innovations and that ultimately fail to deliver on their promises of medical progress for patients. It was indeed pointed out that 1) the results of early trials always overestimate the observed benefits [18–21] and 2) Innovation is different to research and practice and that innovation is not always successful and its dissemination or adoption is sometimes premature. I. Kerridge warned that « innovation (and research) can also be rubbish, needlessly repetitive, wasteful and harmful”, “Innovation may also not be desired (always and at all cost) ie it may not be worth the collapse of PBAC standards of evidence, or the degradation of the public hospital system, or the collapse of the PBS” and he concluded that « An innovation can even be unethical ». Beyond the professional (and personal) virtues often highlighted to define the criteria for ethical research and innovation (kindness, benefits, harm prevention, integrity, etc.), Kerridge brought to light and facilitated a discussion of criteria that are just as important but all too often overlooked [22]: Prudence (sagacity), Intellectual humility, and Presence. He did, in fact, point out that presence is a cornerstone of surgical and medical ethics, and he restated Little’s definition : « Presence is a virtue and a duty, Presence is real, physical, cognitive, relational and engaged, it persists irrespective of the outcome, bears witness and is forced to learn » [23].

## 4. Conclusion

This international study, conducted in 37 countries between 2000 and 2023, highlights significant socioeconomic inequalities in cancer mortality among adolescents and young adults. Countries with the highest levels of human development, health care coverage, and resources generally had the lowest mortality rates. Descriptive, temporal, and multivariate analyses all point to a significant association between socioeconomic determinants and mortality in this population.

The results underscore the need to strengthen strategies for better professionals’ training (i.e., raising awareness of the specific characteristics of the AYA age group), early diagnosis, and equitable access to cancer care among adolescents and young adults (AYAs). They also highlight the importance of further research into the determinants of the observed inequalities, in order to better understand the underlying mechanisms and guide public health policies targeting this population. These findings underscore the importance of integrating socioeconomic determinants into international strategies to combat cancer among adolescents and young adults. Subsequent country-specific studies [24,25] would allow for a comparison of their effectiveness with that observed among adults and older adults.

## Declaration statement

Because the study is not a clinical trial:

· Clinical trial number is not applicable.

· Ethical approval is not applicable

· Consent to participate is not applicable

· Consent to publish is not applicable

## Data Availability Statement

Epidemiological data have been extracted from Global Burden of Disease database: https://www.healthdata.org/research-analysis/gbd. Socioeconomic indicators have been obtained from World Bank: https://databank.worldbank.org/Human-development-index/id/363d401b, and from United Nations Development Programme: https://www.undp.org/.

## Annexes

### A) List of the studied countries

| **Weak HDI** | **Middle HDI** | **High HDI** | **Very high HDI** |
|---|---|---|---|
| Pakistan | Ivory Coast | Botswana | Argentina |
| Senegal | India | Brazil | Australia |
|  | Kenya | China | Canada |
|  | Nigeria | Egypt | Chile |
|  | Togo | Indonesia | France |
|  |  | Iran | Germany |
|  |  | Marocco | Italy |
|  |  | Mexico | Japan |
|  |  | Peru | Netherlands |
|  |  | South Africa | Norway |
|  |  | Thailand | Poland |
|  |  |  | Russia |
|  |  |  | South Korea |
|  |  |  | Spain |
|  |  |  | Sweden |
|  |  |  | Switzerland |
|  |  |  | Turkey |
|  |  |  | United Kingdom |
|  |  |  | USA |

### B) Map of most common cancers

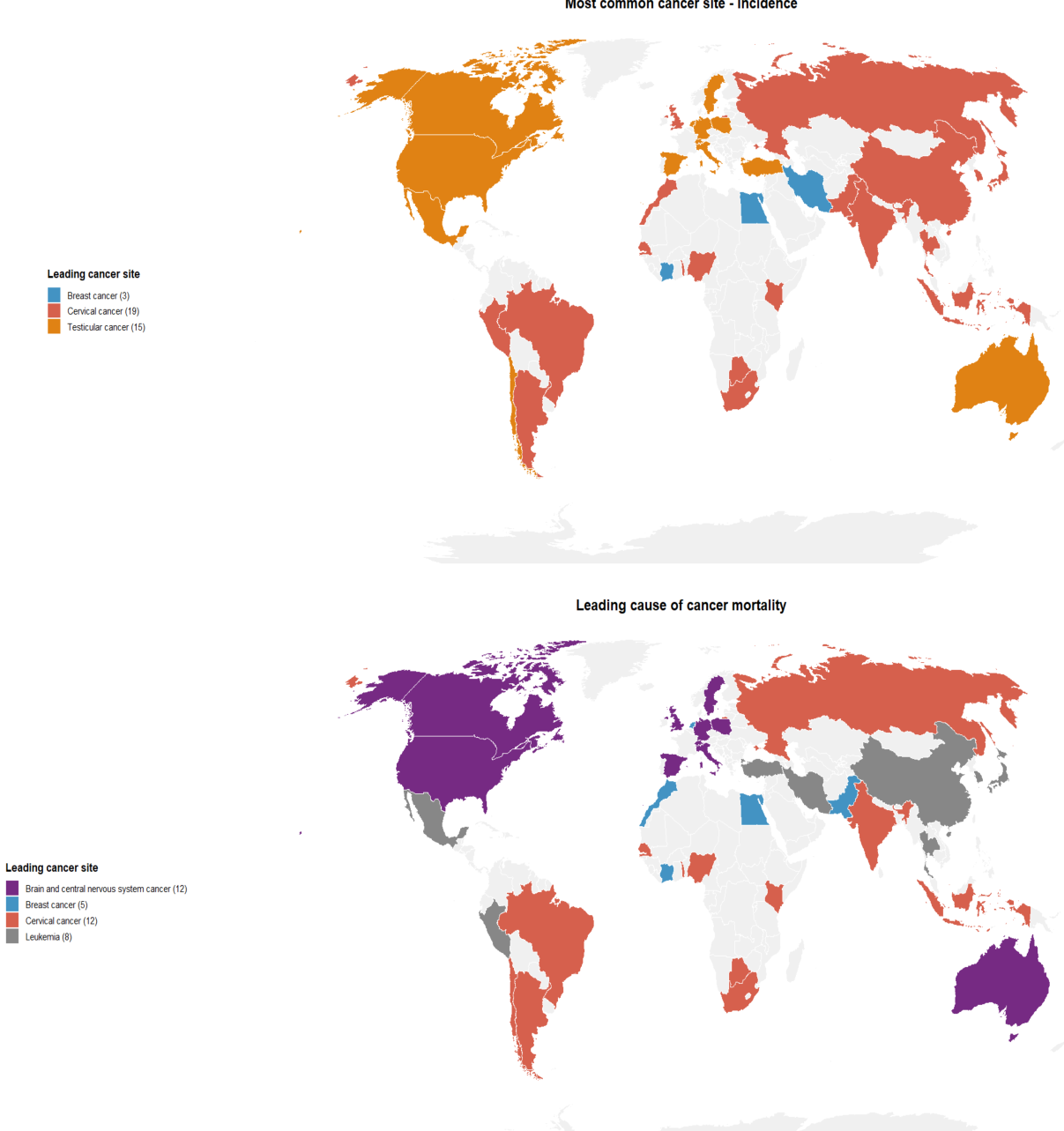

### C) List of Abbreviations

AAPC: Average Annual Percent Change
APC: Annual Percent Change
PCA: Principal Component Analysis
AYA: Adolescents & Young Adults
ASIR: Age-Standardized Incidence Rate
ASMR: Age-Standardized Mortality Rate
EAPC: Estimated Annual Percent Change
GBD: Global Burden of Disease
HCPC: Hierarchical Clustering on Principal Components
HDI: Human Development Index
IHME: Institute for Health Metrics and Evaluation
MIR: Mortality-to-Incidence Rate
UHC: Universal Health Coverage
UNDP: United Nations Development Programme
WHO: World Health Organization

### D) Average annual percent changes (AAPC), 2000-2023

| **Cancer site / Sex** | **AAPC (%)** | **95% CI** | **p-value** |
|---|---|---|---|
| Total cancers (Female) | -0.28 | [-0.41, -0.15] | <0.001 |
| Total cancers (Male) | -0.88 | [-0.97, -0.80] | <0.001 |
| Breast cancer (Female) | 0.93 | [0.79, 1.07] | <0.001 |
| Breast cancer (Male) | 0.00 | [-0.25, 0.20] | 0.906 |
| Cervical cancer (Female) | -0.46 | [-0.82, -0.21] | 0.001 |
| Ovarian cancer (Female) | 0.51 | [0.41, 0.62] | <0.001 |
| Thyroid cancer (Female) | 0.57 | [0.45, 0.69] | <0.001 |
| Thyroid cancer (Male) | -0.23 | [-0.39, -0.08] | 0.002 |
| Brain & NS cancer (Female) | -0.32 | [-0.46, -0.21] | <0.001 |
| Brain & NS cancer (Male) | -0.24 | [-0.31, -0.18] | <0.001 |
| Colorectal cancer (Female) | 0.23 | [0.11, 0.35] | <0.001 |
| Colorectal cancer (Male) | -0.02 | [-0.16, 0.08] | 0.637 |
| Testicular cancer (Male) | -0.27 | [-0.40, -0.13] | <0.001 |
| Leukemia (Female) | -1.29 | [-1.38, -1.22] | <0.001 |
| Leukemia (Male) | -1.03 | [-1.09, -0.99] | <0.001 |
| Hodgkin lymphoma (Female) | -2.37 | [-2.48, -2.24] | <0.001 |
| Hodgkin lymphoma (Male) | -2.03 | [-2.11, -1.95] | <0.001 |
| Non-Hodgkin lymphoma (Female) | -1.11 | [-1.23, -0.98] | <0.001 |
| Non-Hodgkin lymphoma (Male) | -1.03 | [-1.11, -0.94] | <0.001 |
| Sarcomas (Female) | -0.20 | [-0.30, -0.09] | 0.002 |
| Sarcomas (Male) | -0.11 | [-0.19, -0.00] | 0.048 |

### E) Joinpoint-identified period-specific trends (APC), 2000-2023

| **Cancer site** | **Period** | **APC (%)** | **95% CI** | **p-value** |
|---|---|---|---|---|
| Ovarian cancer | 2000-2006 | -1.81 | [-2.71 ; -1.30] | <0.001 |
| Ovarian cancer | 2006-2014 | 0.04 | [-0.43 ; 0.64] | 0.848 |
| Ovarian cancer | 2014-2023 | 2.53 | [2.23 ; 2.93] | <0.001 |
| Thyroid cancer (Female) | 2000-2006 | -1.90 | [-3.00 ; -1.32] | <0.001 |
| Thyroid cancer (Female) | 2006-2012 | 0.10 | [-0.79 ; 1.46] | 0.735 |
| Thyroid cancer (Female) | 2012-2023 | 2.20 | [1.95 ; 2.66] | <0.001 |
| Thyroid cancer (Male) | 2000-2014 | -0.69 | [-1.18 ; -0.45] | <0.001 |
| Thyroid cancer (Male) | 2014-2023 | 0.49 | [0.02 ; 1.58] | 0.040 |
| Brain & NS cancer (Female) | 2000-2007 | -1.02 | [-1.85 ; -0.57] | 0.024 |
| Brain & NS cancer (Female) | 2007-2019 | -0.50 | [-0.95 ; 0.02] | 0.060 |
| Brain & NS cancer (Female) | 2019-2023 | 1.47 | [0.52 ; 2.52] | 0.002 |
| Brain & NS cancer (Male) | 2000-2006 | -0.90 | [-1.50 ; -0.63] | <0.001 |
| Brain & NS cancer (Male) | 2006-2020 | -0.23 | [-0.35 ; -0.12] | 0.004 |
| Brain & NS cancer (Male) | 2020-2023 | 1.10 | [0.37 ; 1.46] | <0.001 |
| Cervical cancer | 2000-2007 | -4.52 | [-6.46 ; -3.52] | <0.001 |
| Cervical cancer | 2007-2020 | 0.79 | [-2.20 ; 1.27] | 0.222 |
| Cervical cancer | 2020-2023 | 3.95 | [1.12 ; 5.77] | <0.001 |
| Colorectal cancer (Female) | 2000-2006 | -2.34 | [-3.40 ; -1.74] | <0.001 |
| Colorectal cancer (Female) | 2006-2014 | 0.16 | [-0.54 ; 0.79] | 0.569 |
| Colorectal cancer (Female) | 2014-2023 | 2.05 | [1.70 ; 2.64] | <0.001 |
| Colorectal cancer (Male) | 2000-2009 | -1.57 | [-2.16 ; -1.25] | <0.001 |
| Colorectal cancer (Male) | 2009-2019 | 0.42 | [-0.38 ; 0.79] | 0.145 |
| Colorectal cancer (Male) | 2019-2023 | 2.41 | [1.32 ; 3.53] | <0.001 |
| Breast cancer (Female) | 2000-2006 | -2.76 | [-3.89 ; -2.05] | <0.001 |
| Breast cancer (Female) | 2006-2013 | 0.53 | [-0.38 ; 1.45] | 0.196 |
| Breast cancer (Female) | 2013-2023 | 3.50 | [3.13 ; 3.99] | <0.001 |
| Breast cancer (Male) | 2000-2004 | -1.89 | [-3.14 ; -0.49] | 0.004 |
| Breast cancer (Male) | 2004-2020 | -0.20 | [-0.53 ; 0.29] | 0.258 |
| Breast cancer (Male) | 2020-2023 | 3.63 | [1.29 ; 4.68] | 0.002 |
| Testicular cancer | 2000-2005 | -2.35 | [-3.69 ; -1.55] | <0.001 |
| Testicular cancer | 2005-2014 | -0.45 | [-0.98 ; 0.19] | 0.118 |
| Testicular cancer | 2014-2023 | 1.10 | [0.75 ; 1.83] | 0.002 |
| Leukemia (Female) | 2000-2012 | -2.01 | [-2.34 ; -1.87] | <0.001 |
| Leukemia (Female) | 2012-2019 | -1.17 | [-1.72 ; -0.67] | <0.001 |
| Leukemia (Female) | 2019-2023 | 0.68 | [-0.02 ; 1.56] | 0.055 |
| Leukemia (Male) | 2000-2011 | -1.82 | [-1.94 ; -1.72] | <0.001 |
| Leukemia (Male) | 2011-2019 | -0.80 | [-1.04 ; -0.58] | <0.001 |
| Leukemia (Male) | 2019-2023 | 0.68 | [0.24 ; 1.29] | 0.003 |
| Hodgkin lymphoma (Female) | 2000-2006 | -3.11 | [-4.30 ; -2.51] | <0.001 |
| Hodgkin lymphoma (Female) | 2006-2023 | -2.11 | [-2.23 ; -1.82] | <0.001 |
| Hodgkin lymphoma (Male) | 2000-2023 | -2.03 | [-2.11 ; -1.95] | <0.001 |
| Non-Hodgkin lymphoma (Female) | 2000-2007 | -2.60 | [-3.38 ; -2.04] | <0.001 |
| Non-Hodgkin lymphoma (Female) | 2007-2023 | -0.44 | [-0.62 ; -0.25] | <0.001 |
| Non-Hodgkin lymphoma (Male) | 2000-2005 | -2.59 | [-3.51 ; -2.05] | <0.001 |
| Non-Hodgkin lymphoma (Male) | 2005-2013 | -1.08 | [-1.55 ; -0.64] | <0.001 |
| Non-Hodgkin lymphoma (Male) | 2013-2023 | -0.20 | [-0.39 ; 0.25] | 0.239 |
| Sarcomas (Female) | 2000-2006 | -1.80 | [-2.89 ; -1.24] | <0.001 |
| Sarcomas (Female) | 2006-2014 | -0.43 | [-0.98 ; 0.45] | 0.208 |
| Sarcomas (Female) | 2014-2023 | 1.08 | [0.79 ; 1.85] | 0.005 |
| Sarcomas (Male) | 2000-2004 | -2.17 | [-3.27 ; -1.26] | <0.001 |
| Sarcomas (Male) | 2004-2023 | 0.32 | [0.23 ; 0.43] | <0.001 |
| Total cancers (Female) | 2000-2006 | -2.96 | [-4.04 ; -2.31] | <0.001 |
| Total cancers (Female) | 2006-2014 | -0.43 | [-1.10 ; 0.24] | 0.174 |
| Total cancers (Female) | 2014-2023 | 1.68 | [1.30 ; 2.27] | <0.001 |
| Total cancers (Male) | 2000-2006 | -2.56 | [-3.14 ; -2.17] | <0.001 |
| Total cancers (Male) | 2006-2019 | -0.82 | [-0.99 ; -0.67] | <0.001 |
| Total cancers (Male) | 2019-2023 | 1.52 | [0.83 ; 2.52] | <0.001 |

